# E-cigarette, combustible, and smokeless tobacco product use combinations among youth in the United States, 2014-2019

**DOI:** 10.1101/2020.06.11.20112581

**Authors:** Jamie Tam

## Abstract

**Introduction:** Youth e-cigarette use has been rising, however U.S. prevalence data are generally reported without disaggregating by individuals’ use of other tobacco products. It is not clear how the proportion of youth e-cigarette users naïve to all combustible tobacco is changing.

**Methods:** Annual prevalence estimates of ever and current (defined as past 30-day use) tobacco use prevalence by school type are reported using the 2014-2019 National Youth Tobacco Surveys (NYTS) with mutually exclusive categories of e-cigarette, smokeless tobacco, and/or combustible tobacco product use. T-tests were used to compare annual estimates with the preceding year. The annual percent change (APC) for each category from 2014-2018 were analyzed using JoinPoint regression. Data for 2019 were reported separately due to the change in survey format from paper to electronic.

**Results:** Current use of only e-cigarettes among HS students who never used combustible tobacco increased significantly from 2014-2018 (APC = +42.4%, 95% CI: 0.7, 101.3); by 2019, prevalence peaked at 9.2% (95% CI: 8.2, 10.2) among never combustible users and 8.3% (95% CI: 7.3, 9.3) among former combustible users. This coincided with significant declines in use of only combustible tobacco (APC=-14.5%, 95% CI: −18.3, −10.5).

**Conclusions:** Use of only e-cigarettes among US youth with no history of combustible tobacco use has increased substantially over time, even as combustible tobacco use continues to plummet. Of the 17.5% (95% CI: 15.7, 19.0) of HS students who currently used only e-cigarettes (but not other tobacco) in 2019, more than half have no history of combustible tobacco use.

## INTRODUCTION

Each year, US health authorities report the latest estimates of youth tobacco use prevalence across different product types including cigarettes, cigars, smokeless tobacco, e-cigarettes, hookahs, pipe tobacco, and bidis.^1^ These reports show that e-cigarettes have been the most commonly used tobacco product among high school and middle school students since 2014. The National Youth Tobacco Surveys showed dramatic increases in e-cigarette use from 2017 to and this trend continued in 2019 when 27.5% high school students reported past 30-day use, with JUUL being the most commonly used brand.^2,3^ These data prompted large-scale efforts by health authorities to curb the youth e-cigarette “epidemic” across the country.^4-6^

Yet among these youth e-cigarette users, it is unknown what proportion of them are using only e-cigarettes (but not other tobacco products) and importantly, how this has changed over time. Tobacco product user groups overlap, and individuals who use e-cigarettes could also be using combustible tobacco, smokeless tobacco or both. For example, research shows that e-cigarette use has occurred predominately among youth who are already smoking cigarettes.^7-9^ Furthermore, previous data indicate that susceptibility to trying e-cigarettes is higher for youth who have previously ever tried using other tobacco products.^10^ It is therefore important to understand whether e-cigarette users have histories of using other tobacco products as well, especially combustible tobacco.

Without disaggregating each tobacco use group by individuals’ current or past use of other tobacco products, it is not possible to evaluate how e-cigarettes are being used in combination or isolation over time. This study estimates the annual proportion of U.S. youth who report ever and current use of any tobacco product across mutually exclusive tobacco product combinations with respect to e-cigarettes, combustible tobacco, and smokeless tobacco products.

## METHODS

### Data source

The National Youth Tobacco Survey (NYTS) is a nationally-representative cross-sectional survey of high school (HS) and middle school (MS) students conducted annually across private and public schools in the United States. From 2014-2018, the NYTS was administered as a pencil-and-paper survey with the following sample sizes (response rates) in 2014=22,007 (73.3%), 2015=17,711 (63.4%), 2016=20,675 (71.6%), 2017=17,872 (68.1%), and 2018=20,189 (68.2%). In 2019, the NYTS was administered electronically for the first time with sample size (response rate) of 19,018 (66.3%).

### Measures

Ever use of e-cigarettes was defined based on a yes or no response to: “*Have you ever used an e-cigarette, even once or twice?”* In 2014, students were asked if they had “*ever tried an electronic cigarette such as Blu, 21st Century Smoke, or NJOY.”* In 2015, students were asked if they had ever used an e-cigarette, “*even once or twice”* and were provided with a list of brands such as “*NJOY, Blu, VUSE, MarkTen, Finiti, Starbuzz, and Fantasia.”* The latter three brands were removed as examples in 2016 and replaced with “*Logic, Vapin Plus, eGo, and Halo”*. JUUL was not included as a brand example in the survey until 2019.

Ever use of combustible tobacco was defined as ever use of cigarettes (including roll-your-own), cigars (cigars, cigarillos, little cigars), pipe tobacco, hookah, or bidis—even “*one or two puffs”*. Ever smokeless tobacco (ST) use was defined as ever use of chewing tobacco, snuff, dip, snus, or dissolvables, “*even just one time”* or “*even just a small amount”*. Ever use of any tobacco product was defined as ever use of any of the aforementioned products including e-cigarettes, combustible tobacco, and/or ST.

Current use of a tobacco product was defined as any use of that product type within the past 30 days. Questions were worded for cigarettes as: “*During the past 30 days, on how many days did you smoke cigarettes?”;* for chewing tobacco, snuff, or dip as: “*During the past 30 days, on how many days did you use chewing tobacco, snuff, or dip?”;* for e-cigarettes as: “*During the past 30 days, on how many days did you use e-cigarettes?*”; and for hookah as: “*During the past 30 days, on how many days did you smoke tobacco in a hookah or waterpipe?”* Finally, snus, dissolvables, bidis, pipe tobacco, and roll-your-own cigarettes were each listed as possible responses to: “*In the past 30 days, which of the following products have you used on at least one day? (Select one or more)”* Current use of any tobacco product was defined as use of e-cigarettes, combustible tobacco, and/or ST in the past 30 days.

### Analyses

First, the proportion of youth that reported ever using tobacco were assessed across six mutually exclusive tobacco use categories and by school type:

1. E-cigarettes only
2. ST only OR ST and e-cigarettes;
3. Combustible tobacco only;
4. Combustible tobacco and e-cigarettes;
5. Combustible and smokeless tobacco;
6. Combustible and smokeless tobacco, and e-cigarettes.

For instance, those who reported ever using e-cigarettes only (category #1 above) also reported never using combustible tobacco or ST. Because ‘ST only’ users represent a very small proportion of youth (<1%) and there were no statistically significant differences in this estimate from 2014-2018, this group was combined with ever users of both ST and e-cigarettes (category #2 above).

Next, the percent of students who were current users across each of the above tobacco use categories are estimated. However, one additional modification to the categories listed above was made: current users of e-cigarettes only ware further disaggregated based on history of combustible tobacco use. That is, use of e-cigarettes only among those who have never used combustible tobacco and those who have ever used (but do not currently use) combustible tobacco were assessed separately. The breaks down current users into seven mutually exclusive categories of product use combinations.

To assess the statistical significance of trends in ever or current use from 2014-2018, Joinpoint regression was used to report the annual percent change (APC) in prevalence for each of the above mutually exclusive tobacco use categories.^11^ In addition, t-tests were performed to determine whether changes between two consecutive survey years were significant, such as from 2017 to 2018. Data from 2019 were not included in Joinpoint or t-test analyses because the shift in survey format for paper and pencil to electronic administration did not allow for direct statistical comparison with previous survey years. Consistent with previous reports,^1^ individuals who reported no use of any type of tobacco product were treated as non-users of tobacco. Those with missing data for all tobacco product types were treated as missing observations. This study was conducted in R 4.0.0 using the ‘survey’ package.^12^

## RESULTS

Ever tobacco use prevalence by product categories and school type are shown in Table 1. From 2014-2018, there were no statistically significant trends in the proportion of HS or MS students who reported ever use of any tobacco product. However by 2019, ever use of any tobacco product was higher than all preceding years at 53.3% for HS students and 24.3% for MS students. Ever use of e-cigarettes only increased from 2014-2018, though t-tests show that the year-to-year change for both HS and MS students were only significant from 2014-2015 and 2017-2018. Joinpoint analyses found a significant increasing trend from 2014-2018 for MS students who had ever used only e-cigarettes and not other tobacco products (APC=+13.3%). An increase in e-cigarette use only was observed for HS students, but was not statistically significant (APC=+20.7%). By 2019, ever use of only e-cigarettes was 19.5% among HS students and 9.7% among MS students - much higher than in 2018 (HS: 11.6%; MS: 5.7%). For both HS and MS students, ever use of ST with or without e-cigarettes remained relatively stable at <2% from 2014-2019.

Irrespective of ST or e-cigarette use, ever use of any combustible tobacco among HS students decreased from 41.8% (95% CI: 39.6, 44.0) in 2014 to 32.1% (95% CI: 28.9, 35.3) in 2019, with declines among MS students as well (2014:14.8%, 95% CI: 12.6,17.0; 2019:13.0%, 95% Cl: 11.4,15.0). From 2014-2018, there were significant declines in HS students’ ever use of combustible tobacco only (APC=-16.1%) and of combustible and smokeless tobacco but not e-cigarettes (APC=-16.0%); such declines were not significant among middle school students during this time period. For ever use of both e-cigarettes and combustible tobacco among HS students, there was a significant increase from 2014-2015, followed by significant decreases between sequential survey years through 2017. Ever use of both e-cigarettes and combustible tobacco also reached its highest in 2019, when 16.9% and 6.6% of HS and MS students reported both products’ use.

**Table 1.** Prevalence of ever tobacco use by product type and school level, National Youth Tobacco Survey, 2014-2019

| School level / Tobacco product(s) ever used | 2014 | 2015 | 2016 | 2017 | 2018 | 2019 | APC 2014-2018 |
| --- | --- | --- | --- | --- | --- | --- | --- |
| **High school** | % (95% CI) |  |  |  |  |  |  |
| Any tobacco product | 47.1 (44.5, 49.8) | 50.4 (47.9, 52.9) | 44.6* (41.8, 47.5) | 41.7 (38.2, 45.2) | 46.2* (44.1, 48.3) | 53.3<sup>^</sup> (50.5, 56.1) | -1.7 (-7.2, 4.2) |
| E-cig only<sup>1</sup> | 3.7 (2.9, 4.8) | 7.4* (6.6, 8.4) | 6.9 (5.9, 8) | 8.0 (7.1, 9) | 11.6* (10.3, 13) | 19.5<sup>^</sup> (17.9, 21.1) | 20.7 (-1.5, 47.8) |
| ST only (OR ST + e-cig)<sup>2</sup> | 1.5 (1.1, 2.0) | 1.4 (1.1, 1.8) | 1.2 (1.0, 1.5) | 1.4 (1.0, 1.9) | 1.4 (1.1, 1.7) | 1.7<sup>^</sup> (1.3, 2.2) | -0.8 (-9.8, 9.2) |
| Combustible only<sup>3</sup> | 15.2 (13.7, 16.8) | 9.6* (8.2, 11.1) | 10.0 (8.9, 11.1) | 8.5 (7.5, 9.6) | 7.3 (6.3, 8.5) | 4.5<sup>^</sup> (3.5, 5.5) | -16.1* (-25.3, -5.7) |
| Combustible + e-cig<sup>4</sup> | 14.5 (13.1, 16.0) | 19.9* (18.5, 21.5) | 13.8* (12.3, 15.3) | 11.7* (10.5, 13) | 13.2 (12.1, 14.5) | 16.9<sup>^</sup> (15.7, 18.1) | -7.8 (-24.8, 12.9) |
| Combustible + ST<sup>5</sup> | 3.1 (2.4, 4.0) | 1.9* (1.4, 2.6) | 2.0 (1.6, 2.5) | 2.0 (1.5, 2.6) | 1.3 (1, 1.8) | 1.0<sup>^</sup> (0.7, 1.4) | -16.0* (-29.1, -0.5) |
| Combustible + ST + e-cig<sup>6</sup> | 8.3 (7, 9.7) | 9.1 (7.6, 10.9) | 9.5 (8.0, 11.2) | 8.9 (7.2, 11.1) | 9.9 (8.7, 11.3) | 9.7<sup>^</sup> (7.3, 12.1) | 3.8 (-0.2, 7.8) |
| **Middle school** | % (95% CI) |  |  |  |  |  |  |
| Any tobacco product | 19.1 (16.6, 21.7) | 19.4 (17.0, 22.0) | 19.5 (17.5, 21.7) | 16.2* (14.7, 17.8) | 18.0 (16.3, 19.9) | 24.3<sup>^</sup> (22.3, 26.3) | -3.1 (-10.8, 5.2) |
| E-cig only<sup>1</sup> | 2.9 (2.3, 3.5) | 4.4* (3.8, 5.1) | 4.5 (3.9, 5.2) | 4.6 (3.9, 5.6) | 5.7* (5.1, 6.4) | 9.7<sup>^</sup> (8.7, 10.8) | 13.3* (1.2, 26.9) |
| ST only (OR ST + e-cig)<sup>2</sup> | 1.1 (0.7, 1.6) | 1.4 (0.9, 2) | 1.3 (0.9, 1.9) | 1.1 (0.7, 1.5) | 1.4 (1.1, 1.8) | 1.6<sup>^</sup> (1.1, 2.0) | 2.9 (-9.5, 16.8) |
| Combustible only<sup>3</sup> | 6.5 (5.4, 7.8) | 4.0 (3.3, 4.7) | 4.3 (3.7, 5.1) | 3.5* (3.0, 4.1) | 3.8 (3.2, 4.6) | 2.9<sup>^</sup> (2.2, 3.5) | -11.3 (-25.7, 5.7) |
| Combustible + e-cig<sup>4</sup> | 4.5 (3.8, 5.3) | 6.2 (5.4, 7.2) | 5.0* (4.3, 5.7) | 3.4* (2.9, 4) | 3.4 (2.8, 4) | 6.6<sup>^</sup> (5.7, 7.5) | -11.5 (-29.7, 11.4) |
| Combustible + ST<sup>5</sup> | 1.3 (0.9, 1.9) | 0.6 (0.4, 0.9) | 1.2* (0.8, 1.9) | 0.9 (0.5, 1.4) | 0.8 (0.6, 1.1) | 0.6<sup>^</sup> (0.4, 0.9) | -6.8 (-29.1, 22.7) |
| Combustible + ST + e-cig<sup>6</sup> | 2.2 (1.5, 3.2) | 2.2 (1.7, 2.9) | 2.7 (2.1, 3.5) | 2.1 (1.7, 2.7) | 2.0 (1.6, 2.6) | 2.8<sup>^</sup> (2.2, 3.4) | -3.5 (-16.6, 11.7) |

Abbreviations: CI = confidence interval; E-cig = e-cigarette; ST = smokeless tobacco; APC = Annual percent change.
\*p-value <0.05 in t-tests comparing prevalence of ever use with previous survey year or in APC Joinpoint analyses for 2014-2018.
<sup>^</sup>Data from 2019 are not directly comparable with estimates in previous years due to changes in the NYTS format from paper to electronic.
<sup>1</sup>Ever used e-cigarettes only, and never used combustible or smokeless tobacco. <sup>2</sup>Ever used smokeless tobacco (including chewing tobacco, snuff, dip, snus or dissolvables), but never used combustible tobacco. Includes individuals who may have ever used e-cigarettes. <sup>3</sup>Ever used combustible tobacco (cigarettes, cigars, cigarillos, little cigars, pipe tobacco, hookah, and/or bidis), but never used e-cigarettes or smokeless tobacco. <sup>4</sup>Ever used combustible tobacco and e-cigarettes, but never used smokeless tobacco. <sup>5</sup>Ever used combustible and smokeless tobacco, but never used e-cigarettes. <sup>6</sup>Ever used combustible tobacco, smokeless tobacco, and e-cigarettes.

Current tobacco use prevalence by product categories and school type are reported in Table 2. Trends in past 30-day use of any tobacco product were non-significant for HS and MS students over the 2014-2018 time period. However by 2019, use of any tobacco product peaked at 31.2% and 12.5% for HS and MS students respectively. From 2014-2018, significant increases occurred in use of only e-cigarettes among HS students who had never used combustible tobacco (APC=+42.4%). In 2018,11.4% (95% CI: 9.7,13.0) of HS students were current users of only e-cigarettes, half of whom had never used combustible tobacco (5.7%) compared to those who had (5.4%). By 2019,17.5% (95% CI: 15.7,19.0) of HS students used only e-cigarettes; slightly more of whom had never used combustible tobacco (9.2%) compared to those who had (8.3%); for MS students, 6.9% (95% CI: 6.0, 8.0) used only e-cigarettes, a larger proportion of whom had no history of combustible tobacco use (4.5%). For both HS and MS students, current use of ST with or without e-cigarettes remained relatively stable from 2014-2019.

Irrespective of ST or e-cigarette use, current use of any combustible tobacco among HS students decreased from 17.9% (95% CI: 16.4,19.0) in 2014 to 12.0% (95% CI: 10.5,13.0) in 2019. From 2014-2018, there were significant decreases in the use of combustible tobacco products only among HS students (APC=-14.5%) and non-significant decreases among MS students (APC=-13.9%). In 2019, use of combustible tobacco and e-cigarettes among HS and MS students was 6.3% and 2.5% respectively. Among HS students in 2019, use of only combustible tobacco dropped to its lowest at 2.6% and use of combustible tobacco with ST to 0.4%.

**Table 2.**
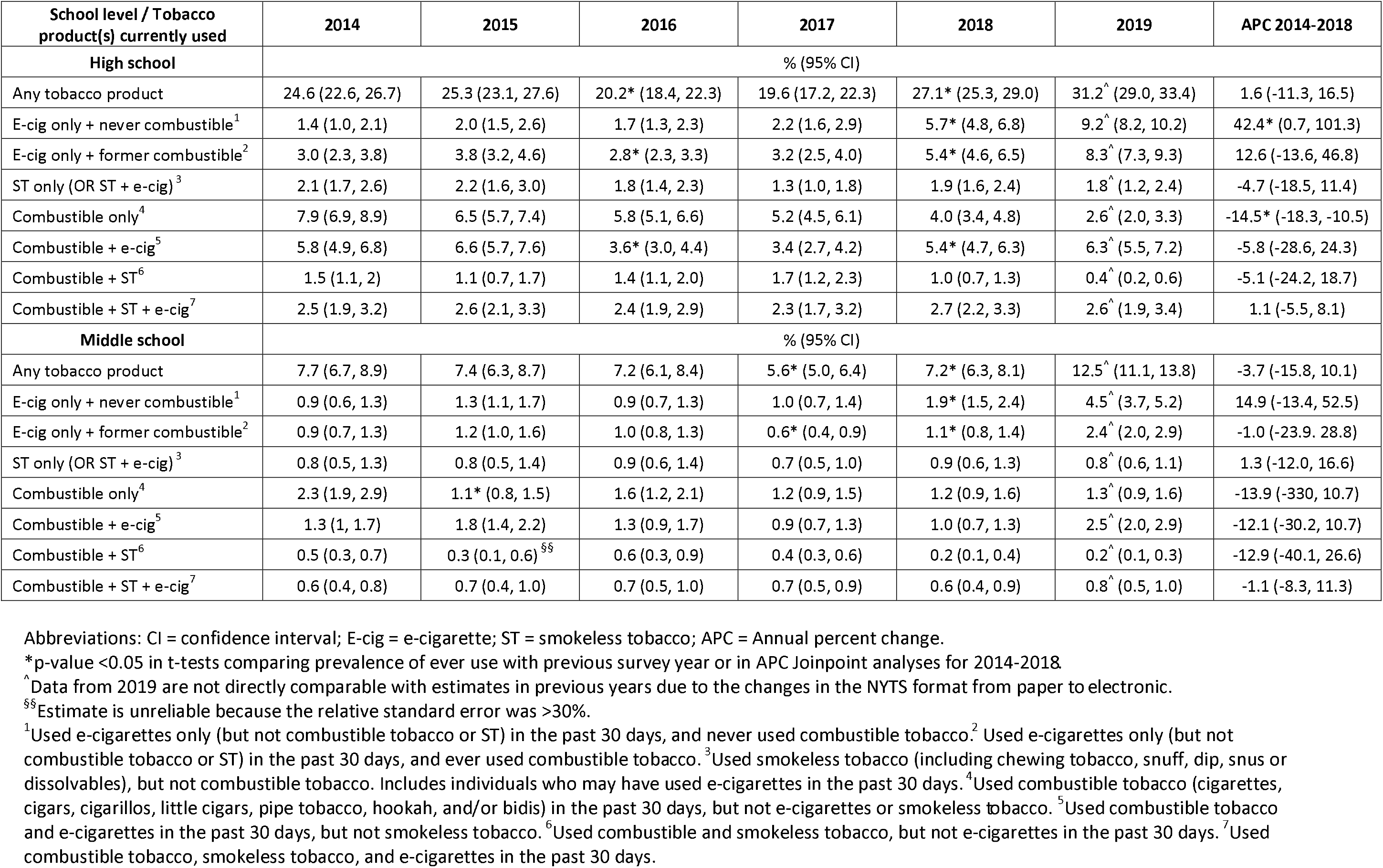
Prevalence of current tobacco use by product type and school level, National Youth Tobacco Survey, 2014-2019

| School level / Tobacco product(s) currently used | 2014 | 2015 | 2016 | 2017 | 2018 | 2019 | APC 2014-2018 |
| --- | --- | --- | --- | --- | --- | --- | --- |
| **High school** | % (95% CI) |  |  |  |  |  |  |
| Any tobacco product | 24.6 (22.6, 26.7) | 25.3 (23.1, 27.6) | 20.2* (18.4, 22.3) | 19.6 (17.2, 22.3) | 27.1* (25.3, 29.0) | 31.2<sup>^</sup> (29.0, 33.4) | 1.6 (-11.3, 16.5) |
| E-cig only + never combustible<sup>1</sup> | 1.4 (1.0, 2.1) | 2.0 (1.5, 2.6) | 1.7 (1.3, 2.3) | 2.2 (1.6, 2.9) | 5.7* (4.8, 6.8) | 9.2<sup>^</sup> (8.2, 10.2) | 42.4* (0.7, 101.3) |
| E-cig only + former combustible<sup>2</sup> | 3.0 (2.3, 3.8) | 3.8 (3.2, 4.6) | 2.8* (2.3, 3.3) | 3.2 (2.5, 4.0) | 5.4* (4.6, 6.5) | 8.3<sup>^</sup> (7.3, 9.3) | 12.6 (-13.6, 46.8) |
| ST only (OR ST + e-cig)<sup>3</sup> | 2.1 (1.7, 2.6) | 2.2 (1.6, 3.0) | 1.8 (1.4, 2.3) | 1.3 (1.0, 1.8) | 1.9 (1.6, 2.4) | 1.8<sup>^</sup> (1.2, 2.4) | -4.7 (-18.5, 11.4) |
| Combustible only<sup>4</sup> | 7.9 (6.9, 8.9) | 6.5 (5.7, 7.4) | 5.8 (5.1, 6.6) | 5.2 (4.5, 6.1) | 4.0 (3.4, 4.8) | 2.6<sup>^</sup> (2.0, 3.3) | -14.5* (-18.3, -10.5) |
| Combustible + e-cig<sup>5</sup> | 5.8 (4.9, 6.8) | 6.6 (5.7, 7.6) | 3.6* (3.0, 4.4) | 3.4 (2.7, 4.2) | 5.4* (4.7, 6.3) | 6.3<sup>^</sup> (5.5, 7.2) | -5.8 (-28.6, 24.3) |
| Combustible + ST<sup>6</sup> | 1.5 (1.1, 2) | 1.1 (0.7, 1.7) | 1.4 (1.1, 2.0) | 1.7 (1.2, 2.3) | 1.0 (0.7, 1.3) | 0.4<sup>^</sup> (0.2, 0.6) | -5.1 (-24.2, 18.7) |
| Combustible + ST + e-cig<sup>7</sup> | 2.5 (1.9, 3.2) | 2.6 (2.1, 3.3) | 2.4 (1.9, 2.9) | 2.3 (1.7, 3.2) | 2.7 (2.2, 3.3) | 2.6<sup>^</sup> (1.9, 3.4) | 1.1 (-5.5, 8.1) |
| **Middle school** | % (95% CI) |  |  |  |  |  |  |
| Any tobacco product | 7.7 (6.7, 8.9) | 7.4 (6.3, 8.7) | 7.2 (6.1, 8.4) | 5.6* (5.0, 6.4) | 7.2* (6.3, 8.1) | 12.5<sup>^</sup> (11.1, 13.8) | -3.7 (-15.8, 10.1) |
| E-cig only + never combustible<sup>1</sup> | 0.9 (0.6, 1.3) | 1.3 (1.1, 1.7) | 0.9 (0.7, 1.3) | 1.0 (0.7, 1.4) | 1.9* (1.5, 2.4) | 4.5<sup>^</sup> (3.7, 5.2) | 14.9 (-13.4, 52.5) |
| E-cig only + former combustible<sup>2</sup> | 0.9 (0.7, 1.3) | 1.2 (1.0, 1.6) | 1.0 (0.8, 1.3) | 0.6* (0.4, 0.9) | 1.1* (0.8, 1.4) | 2.4<sup>^</sup> (2.0, 2.9) | -1.0 (-23.9, 28.8) |
| ST only (OR ST + e-cig)<sup>3</sup> | 0.8 (0.5, 1.3) | 0.8 (0.5, 1.4) | 0.9 (0.6, 1.4) | 0.7 (0.5, 1.0) | 0.9 (0.6, 1.3) | 0.8<sup>^</sup> (0.6, 1.1) | 1.3 (-12.0, 16.6) |
| Combustible only<sup>4</sup> | 2.3 (1.9, 2.9) | 1.1* (0.8, 1.5) | 1.6 (1.2, 2.1) | 1.2 (0.9, 1.5) | 1.2 (0.9, 1.6) | 1.3<sup>^</sup> (0.9, 1.6) | -13.9 (-33.0, 10.7) |
| Combustible + e-cig<sup>5</sup> | 1.3 (1, 1.7) | 1.8 (1.4, 2.2) | 1.3 (0.9, 1.7) | 0.9 (0.7, 1.3) | 1.0 (0.7, 1.3) | 2.5<sup>^</sup> (2.0, 2.9) | -12.1 (-30.2, 10.7) |
| Combustible + ST<sup>6</sup> | 0.5 (0.3, 0.7) | 0.3 (0.1, 0.6)<sup>§§</sup> | 0.6 (0.3, 0.9) | 0.4 (0.3, 0.6) | 0.2 (0.1, 0.4) | 0.2<sup>^</sup> (0.1, 0.3) | -12.9 (-40.1, 26.6) |
| Combustible + ST + e-cig<sup>7</sup> | 0.6 (0.4, 0.8) | 0.7 (0.4, 1.0) | 0.7 (0.5, 1.0) | 0.7 (0.5, 0.9) | 0.6 (0.4, 0.9) | 0.8<sup>^</sup> (0.5, 1.0) | -1.1 (-8.3, 11.3) |

Abbreviations: CI = confidence interval; E-cig = e-cigarette; ST = smokeless tobacco; APC = Annual percent change.
\*p-value <0.05 in t-tests comparing prevalence of ever use with previous survey year or in APC Joinpoint analyses for 2014-2018.
<sup>^</sup>Data from 2019 are not directly comparable with estimates in previous years due to the changes in the NYTS format from paper to electronic.
<sup>§§</sup>Estimate is unreliable because the relative standard error was >30%.
<sup>1</sup>Used e-cigarettes only (but not combustible tobacco or ST) in the past 30 days, and never used combustible tobacco. <sup>2</sup>Used e-cigarettes only (but not combustible tobacco or ST) in the past 30 days, and ever used combustible tobacco. <sup>3</sup>Used smokeless tobacco (including chewing tobacco, snuff, dip, snus or dissolvables), but not combustible tobacco. Includes individuals who may have used e-cigarettes in the past 30 days. <sup>4</sup>Used combustible tobacco (cigarettes, cigars, cigarillos, little cigars, pipe tobacco, hookah, and/or bidis) in the past 30 days, but not e-cigarettes or smokeless tobacco. <sup>5</sup>Used combustible tobacco and e-cigarettes in the past 30 days, but not smokeless tobacco. <sup>6</sup>Used combustible and smokeless tobacco, but not e-cigarettes in the past 30 days. <sup>7</sup>Used combustible tobacco, smokeless tobacco, and e-cigarettes in the past 30 days.

## DISCUSSION

This analysis examines trends in youth use of different tobacco product combinations since e-cigarettes came to dominate use in 2014, and disaggregates e-cigarette users by their other tobacco product use. Ever use of only e-cigarettes was the primary driver in the recent 2019 increase in ever use of any tobacco product. In 2019,1 in 5 HS students and 1 in 10 MS students reported ever use of only e-cigarettes, but not other products. That year, 17.5% of HS students had only used e-cigarettes in the past month, up from 11.4% in 2018; use appears to be rising in equal measure among those with prior have formerly ever used combustible tobacco, and those with no history of combustible tobacco use.

Though increases in e-cigarette use are concerning, they are occurring simultaneously with declines in combustible tobacco use. Current use of any combustible tobacco product in the past month among HS students dropped substantially from 17.9% in 2014 to 12.0% in 2019. It is encouraging that high school students’ use of combustible tobacco products continues to drop each year.

With new developments in the tobacco marketplace, including the proliferation of pod-based e-cigarettes known to delivery higher levels of nicotine content such as JUUL,^13^ e-cigarette use - and consequently nicotine exposure—could be taking place among youth who have never tried using any tobacco product or who might not otherwise be susceptible to tobacco. On the other hand, e-cigarettes could also be attracting never users who might have eventually gone on to use cigarettes or other combustible products—diverting would-be smokers away from the most dangerous forms of tobacco. Among the 6.9% of MS students who currently used only e-cigarettes, a majority had no experience with combustible tobacco. Other research using longitudinal data sources can determine whether they will eventually go on to use combustible products in the future or not. For decision-makers to understand youth e-cigarette use, reports should provide detailed breakdowns to account for individuals’ use of other tobacco products, especially combustible tobacco.

## Data Availability

All data are publicly available online.

https://www.cdc.gov/tobacco/data_statistics/surveys/nyts/index.htm

## Acknowledgements

This analysis was partially conducted while the author was a Tobacco Regulatory Science Fellow at the US Food and Drug Administration Center for Tobacco Products. She is immensely grateful to Drs. Gabriella M. Anic and Karen Cullen for their expert guidance, feedback, and support of this project from 2018-2019.

